# Comparing the Treatment and Side Effects of Existing Bariatric Surgery Procedures: An Observational Study

**DOI:** 10.64898/2025.12.17.25342515

**Authors:** Qishuo Yin, Jiawei Zhang, Siyu Heng

## Abstract

**Importance:** Bariatric surgery is an established treatment for obesity and its associated comorbidities, including diabetes, hypertension, sleep apnea, and hypercholesterolemia. Despite the widespread adoption of various bariatric procedures, rigorous causal comparisons of their differential effects on treatment outcomes and adverse events remain scarce.

**Objective:** This large-scale observational study aimed to rigorously compare the effects of commonly performed bariatric surgery procedures on both weight loss (effevtiveness) and the risk of postoperative complications.

**Evidence Review:** This study utilized data from the American College of Surgeons Metabolic and Bariatric Surgery Accreditation and Quality Improvement Program (MB-SAQIP) database, encompassing 729,482 cases from 2015 to 2020. With Sleeve Gastrectomy serving as the reference procedure, we assessed the effect of alternative procedures on changes in body mass index (BMI) and the risk of reoperation, readmission, and subsequent interventions. State-of-the-art machine learning-based causal inference techniques, including Causal Forest, Dragonnet, and Double Machine Learning, were employed to conduct robust causal comparisons.

**Findings:** Biliopancreatic Diversion with Duodenal Switch (BPD/DS) demonstrated superior BMI reduction compared with Sleeve Gastrectomy. Roux-en-Y Gastric Bypass (RYGB), Adjustable Gastric Band (AGB, or Band), and Single Anastomosis Duodeno-Ileal Bypass with Sleeve Gastrectomy (SADI-S) were associated with less pronounced BMI decreases relative to Sleeve Gastrectomy. The risk of complications was similar or higher for all other surgical procedures compared with Sleeve Gastrectomy. Importantly, these represent causal effect estimates rather than mere associations, providing clinically actionable evidence for treatment selection. Detailed effect estimates and risk ratios, along with their confidence intervals, are presented in the full text. All our implementations are available at GitHub.

**Conclusions and Relevance:** Our causal estimates–derived from state-of-the-art machine learning methods applied to the largest bariatric surgery registry–provide the first rigorous quantitative evidence supporting current clinical practice guidelines, issued by the American Society for Metabolic and Bariatric Surgery (ASMBS), and enable evidence-based surgical decision-making.

**Key Points:** *Question:* What are the causal effects of the five widely adopted bariatric surgery procedures on weight loss efficacy and postoperative complication risks?

*Findings:* Our causal analysis reveals that Biliopancreatic Diversion with Duodenal Switch (BPD/DS) achieves significantly greater BMI reduction compared with the most widely conducted Sleeve Gastrectomy, but at the cost of substantially elevated complication risks. Our causal analysis results of all five bariatric surgery procedures align with mechanistic understanding and provide quantitative causal estimates rather than associations.

*Meaning:* This represents the first large-scale and comprehensive causal analysis comparing weight loss and adverse event risks across the five most important bariatric surgery procedures, providing rigorous evidence to inform surgical decision-making.

## 1 Introduction

Overweight and obesity represent pressing global public health challenges [Flegal et al., 2012]. Bariatric surgery has emerged as the most effective intervention for severe obesity, demonstrating unparalleled efficacy in achieving sustained weight loss and resolving obesity-related comorbidities, including type 2 diabetes, cardiovascular disease, and mortality [Sjöström et al., 2007, Adams et al., 2007, Schauer et al., 2017, Courcoulas et al., 2018]. Utilization has increased substantially, with over 252,000 procedures in 2018 in the United States [English et al., 2020], reflecting recognition of surgery’s long-term durability and economic benefits [Maciejewski et al., 2016]. Bariatric surgery comprises five primary operation options–Sleeve Gastrectomy (restrictive), Roux-en-Y Gastric Bypass (RYGB, restrictive-malabsorptive), Adjustable Gastric Band (restrictive), Biliopancreatic Diversion with Duodenal Switch (BPD/DS, malabsorptive), and Single Anastomosis Duodeno-Ileal Bypass with Sleeve Gastrectomy (SADI-S, malabsorptive)–that differ fundamentally in surgical complexity, weight loss mechanisms, and risk profiles [Mechanick et al., 2008]. Each procedure presents distinct trade-offs between weight loss efficacy and complication risk, making comparative effectiveness assessments critical for evidence-based clinical decision-making [Arterburn et al., 2018]. Despite this clinical importance, existing comparative evidence remains largely associational rather than causal, limiting clinical applicability.

Rigorous causal estimation and inference of different bariatric surgery procedure effects is challenging. Traditional associational studies are insufficient because confounding by indication systematically biases comparisons–patients receiving different procedures differ in baseline characteristics, comorbidities, and disease severity, making observed outcome differences unreliable indicators of true treatment effects [Arterburn et al., 2015]. Clinical decision-making requires causal, not merely associational, evidence [Hernán et al., 2019]. While randomized controlled trials (RCTs) represent the gold standard, existing bariatric RCTs predominantly compare surgery to medical management rather than different surgical procedures [Schauer et al., 2017, Mingrone et al., 2021], and comparative RCTs have small sample sizes and limited scope [Helmiö et al., 2014]. Furthermore, strict eligibility criteria limit generalizability, with few real-world patients meeting typical RCT inclusion criteria [Arterburn et al., 2020]. To our knowledge, no prior study has applied rigorous causal inference methods to compare bariatric procedures at this scale. Large-scale registries such as MBSAQIP, when paired with modern causal inference methods, offer a scalable alternative providing causal evidence for clinical decision-making [Courcoulas et al., 2018].

In this work, we apply state-of-the-art machine learning-based causal inference methods to MBSAQIP (*n* = 729,482) to conduct a large-scale observational study of treatment effects and complication risks across bariatric surgery procedures. We evaluate causal impacts on 30-day BMI reduction and postoperative complications (reoperation, readmission, and intervention) using three methods: Causal Forest [Wager and Athey, 2018], Dragonnet [Shi et al., 2019], and Double Machine Learning [Chernozhukov et al., 2018]. These frameworks flexibly model complex covariate-treatment and covariate-outcome relationships. By leveraging state-of-the-art causal inference on MBSAQIP, our work provides, to our knowledge, the first rigorous and comprehensive causal evidence on comparative safety and efficacy of bariatric surgical options, supporting evidence-based treatment decisions and offering guidance for bariatric surgery practitioners.

## 2 Methods

### 2.1 Data Source and Study Population

We analyzed data from the MBSAQIP Data Registry, providing comprehensive national data from accredited bariatric surgery centers with standardized quality assessment Hage et al. [2024]. The registry demonstrates acceptable data completeness and consistency Clapp et al. [2024], systematically collecting detailed preoperative patient characteristics (demographics, comorbidities, anthropometrics, ASA classification) and 30-day postoperative outcomes (BMI changes, complications, readmissions, reoperations), making it well-suited for causal inference studies comparing bariatric procedures.

To rigorously estimate causal treatment effects while minimizing confounding, we included comprehensive preoperative patient characteristics with minimal missing data that are clinically recognized confounders affecting both surgical procedure selection and postoperative outcomes. Our covariates encompassed four major categories: **demographic and baseline factors** (age, sex, race, anthropometric status, functional capacity) influencing surgical risk stratification and procedural selection Santry et al. [2007], Hon et al. [2025]; **cardiovascular and metabolic comorbidities** that determine perioperative risk and often guide surgeons towards less complex procedures for high-risk patients Khan et al. [2013]; **respiratory conditions** influencing anesthetic risk and recovery trajectories Mokhlesi et al. [2013]; and **surgical history and clinical indicators** (previous surgical interventions, medication use, procedural markers) reflecting anatomical considerations and clinical concern for postoperative complications Hackett et al. [2015].

This comprehensive covariate set captures the complexity of surgical decision-making in bariatric practice, where procedure selection is influenced by patient comorbidity burden, anatomical considerations from previous surgeries, and perioperative risk assessment Eisenberg et al. [2006], Yermilov et al. [2009]. By controlling for these preoperative characteristics, our causal inference approach isolates the independent effect of surgical procedure type on outcomes. The complete list of preoperative variables is in Appendix A.

#### 2.1.1 The Five Primary Bariatric Surgery Procedures for Comparisons

We compared five primary bariatric procedures based on their clinical relevance and prevalence in contemporary practice: Sleeve Gastrectomy, Roux-en-Y Gastric Bypass (RYGB), Adjustable Gastric Band (Band), Biliopancreatic Diversion with Duodenal Switch (BPD/DS), and Single Anastomosis Duodeno-Ileal Bypass with Sleeve Gastrectomy (SADI-S). These procedures represent the evolution of bariatric surgery over recent decades and reflect distinct philosophies in balancing weight loss efficacy against surgical complexity and complication risk.

**Sleeve Gastrectomy (Sleeve)** involves removing approximately 80% of the stomach along the greater curvature to create a tubular gastric remnant, reducing stomach capacity while decreasing ghrelin production to achieve mechanical restriction and favorable hormonal changes Gagner et al. [2009], Rosenthal and Panel [2012]. As the most commonly performed bariatric procedure worldwide, accounting for approximately 68% of contemporary operations Angrisani et al. [2015], Schauer et al. [2017], sleeve gastrectomy serves as the baseline against which other procedures are evaluated.

**Roux-en-Y Gastric Bypass (RYGB)** creates a small gastric pouch connected to jejunum, bypassing the majority of the stomach and duodenum through dual anastomosis combining gastric restriction with mild malabsorption and powerful neurohormonal changes, particularly enhanced GLP-1 secretion Mason and Ito [1967], Pories et al. [1995]. RYGB remains important as the former gold standard with over 30 years of outcome data and historically superior metabolic results, particularly for type 2 diabetes remission Courcoulas et al. [2018], Mingrone et al. [2021], Peterli et al. [2018].

**Adjustable Gastric Band (Band)** involves placement of an inflatable silicone band around the proximal stomach to create a small pouch with adjustable restriction, a purely mechanical, reversible approach preserving normal anatomy without resection or intestinal bypass Belachew et al. [1994], O’Brien et al. [2006]. The band merits study as thousands of patients live with bands, making understanding of long-term outcomes and reoperation patterns clinically relevant Lanthaler et al. [2010].

**Biliopancreatic Diversion with Duodenal Switch (BPD/DS)** combines sleeve gastrectomy with extensive intestinal bypass diverting biliopancreatic secretions to the distal ileum, creating significant malabsorption and the most powerful metabolic effects and greatest weight loss among bariatric procedures Marceau et al. [1993], Hess and Hess [1998]. Typically reserved for patients with super obesity or severe metabolic disease Skogar and Sundbom [2017], Prachand et al. [2006], Buchwald et al. [2004], BPD/DS raises questions about optimal balance between efficacy and safety for specific patient populations.

**Single Anastomosis Duodeno-Ileal Bypass with Sleeve Gastrectomy (SADI-S)** is a technical simplification of BPD/DS, replacing dual-anastomosis intestinal reconstruction with a single duodeno-ileal anastomosis after sleeve gastrectomy to reduce operative time and complexity while preserving metabolic benefit Sánchez-Pernaute et al. [2010], Cottam et al. [2016]. SADI-S tests whether technical simplification can maintain metabolic efficacy while reducing operative complexity and surgical risk Topart et al. [2017], reflecting evolution toward simpler procedures in metabolic surgery.
These procedures reflect progression from purely restrictive approaches (Band) toward combined restrictive-malabsorptive techniques (RYGB, BPD/DS), and most recently toward simplified anatomical reconstructions (Sleeve, SADI-S) balancing efficacy with safety. Procedures were classified according to the Code for the Principal Operative Procedure in the MBSAQIP registry.

### 2.2 Outcome Measures

We evaluated postoperative outcomes within 30 days to quantify the risk-benefit trade-off in bariatric surgery: balance between achieving weight loss and minimizing complications. Treatment effectiveness was measured by absolute body mass index (BMI) reduction from preoperative baseline, calculated as the change in BMI units (kg/m^2^) Courcoulas et al. [2013], Schauer et al. [2017]. Surgical risk was quantified by composite safety endpoints of reoperation, hospital readmission, or clinical reintervention, capturing major postoperative morbidity including management of anastomotic leaks, bleeding, bowel obstruction, and other procedure-related complications of Bariatric Surgery [LABS], Hutter et al. [2011].

Understanding how different procedures perform across effectiveness and safety allows surgeons and patients to make informed choices based on individual risk tolerance, obesity severity, and metabolic disease burden Arterburn et al. [2018], Peterli et al. [2018]. The 30-day window aligns with national quality benchmarking programs Dimick et al. [2013], DeMaria et al. [2010] and captures immediate safety profiles and early treatment efficacy indicators.

### 2.3 Statistical Analysis

We conducted an observational study on causal effects of different bariatric surgical procedures relative to sleeve gastrectomy. For weight-loss outcomes, we estimated the average treatment effect (ATE), representing the expected difference in BMI reduction between patients receiving the comparator procedure and those receiving sleeve gastrectomy, after adjusting for measured preoperative covariates influencing both procedure selection and outcomes. For complications, we estimated causal risk ratios (RR), quantifying how many times more (or less) likely complications would occur with the comparator procedure versus sleeve gastrectomy among patients with similar baseline characteristics. Risk ratios greater than 1.0 indicate higher complication rates for the comparator procedure, while ratios less than 1.0 indicate lower rates (formal definitions in Appendix B.1).

We employed machine learning-based causal inference because relationships between patient characteristics and surgical outcomes are inherently nonlinear, with complex interactions between factors such as age, BMI, comorbidities, and metabolic parameters. Traditional parametric methods impose restrictive assumptions that may not reflect biological reality Austin [2011], whereas machine learning approaches flexibly capture nonlinear patterns without requiring pre-specified functional forms Athey and Imbens [2019]. We implemented three approaches: Causal Forest Wager and Athey [2018], using tree-based ensemble learning to estimate heterogeneous treatment effects; Double Machine Learning (DML) Chernozhukov et al. [2018] with three base learners (Random Forest, LightGBM, XGBoost), employing cross-fitting to separately model propensity scores and outcomes; and Dragonnet Shi et al. [2019], a neural network architecture jointly learning both models through shared representations. Concordance across these fundamentally different algorithmic approaches strengthens confidence in findings by reducing sensitivity to specific modeling assumptions Matthay et al. [2020]. For binary complication outcomes, we adapted these methods to estimate risk ratios rather than risk differences, as risk ratios are more clinically interpretable and remain meaningful across populations with different baseline risk levels Zhang and Kai [1998]. Detailed procedures are in Appendix B.3.

## 3 Results

### 3.1 Study Population

Our analysis included 729, 482 bariatric surgery cases from the MBSAQIP registry performed between 2015 and 2020. Descriptive statistics are in Table 1. Sleeve gastrectomy was predominant with 495, 917 cases (68%), followed by RYGB with 198, 100 cases (27%). Less commonly performed procedures include SADI-S with 16, 184 cases (2.2%), Band with 10, 006 cases (1.4%), and BPD/DS with 9, 275 cases (1.3%).

**Table 1:** Descriptive statistics of bariatric surgery data in the MBSAQIP registry (2015-2020). BMI reduction represents the mean change in body mass index units within 30 days post-surgery. Complications include any reoperation, readmission, or intervention within 30 days of the index procedure. Sleeve = Sleeve Gastrectomy; RYGB = Roux-en-Y Gastric Bypass; SADI-S = Single Anastomosis Duodeno-Ileal Bypass with Sleeve Gastrectomy; Band = Adjustable Gastric Band; BPD/DS = Biliopancreatic Diversion with Duodenal Switch.

| Summary \ Surgery |  | Sleeve | RYGB | SADI-S | Band | BPD/DS |
| --- | --- | --- | --- | --- | --- | --- |
| Cases | Number | 495,917 | 198,100 | 16,184 | 10,006 | 9,275 |
|  | Percentage | 67.98% | 27.16% | 2.22% | 1.37% | 1.27% |
| BMI Reduction | Mean | 2.50 | 2.41 | 1.57 | 1.63 | 2.71 |
|  | Standard Error | (0.0032) | (0.0054) | (0.0123) | (0.0163) | (0.0281) |
| Complications | Reoperation | 0.78% | 2.16% | 3.43% | 0.93% | 6.67% |
|  | Readmission | 2.88% | 5.87% | 6.67% | 2.04% | 3.23% |
|  | Intervention | 0.78% | 2.30% | 3.61% | 0.64% | 2.53% |

The average reductions in BMI (i.e., sample means without adjusting for covariates) are reported in Table 1 across the five primary bariatric surgery procedures. BPD/DS demonstrates the greatest mean BMI reduction (2.71 units, *SE* = 0.0281), followed by Sleeve (2.50 units, *SE* = 0.0032) and RYGB (2.41 units, *SE* = 0.0054). Band shows more modest reductions (1.63 units, *SE* = 0.0163), while SADI-S has the smallest reduction (1.57 units, *SE* = 0.0163).

Complication rates (i.e., sample rates without adjusting for covariates) for the five surgery procedures are also reported in Table 1. Complication rates within 30 days of surgery vary substantially across procedures. For reoperations, BPD/DS has the highest rate (6.67%), while Sleeve has the lowest (0.78%). RYGB and SADI-S show intermediate rates (2.16% and 3.43%, respectively), and Band has a relatively low rate (0.93%). Readmission patterns reveal SADI-S with the highest rate (6.67%), followed by RYGB (5.87%), BPD/DS (3.23%), Sleeve (2.88%), and Band (2.04%). Intervention rates follow similar trends, with SADI-S demonstrating the highest rate (3.61%) and Band the lowest (0.64%). BPD/DS, RYGB, and Sleeve show intermediate rates (2.53%, 2.30%, and 0.78%, respectively).

### 3.2 Causal Effect on Postoperative BMI Reduction

Estimates of the average treatment effect (ATE) for 30-day postoperative BMI reduction, comparing each procedure with sleeve gastrectomy, are shown in Table 2 with corresponding 95% confidence intervals. Across all four contrasts (RYGB vs. Sleeve, Band vs. Sleeve, BPD/DS vs. Sleeve, and SADI-S vs. Sleeve), the five ATE estimation and inference approaches yielded directionally consistent results. Although effect sizes varied slightly across methods, the overall pattern of findings was stable, indicating that the estimated causal effects are robust to the choice among the machine-learning-based causal inference methods evaluated.

**Table 2:** Estimation of average treatment effects (ATE) for BMI reduction within 30 days after bariatric surgery compared to Sleeve Gastrectomy (Sleeve). Values represent mean differences in BMI units with 95% confidence intervals in parentheses. Negative values indicate less BMI reduction compared to sleeve gastrectomy; positive values indicate greater reduction. Sleeve = Sleeve Gastrectomy; RYGB = Roux-en-Y Gastric Bypass; SADI-S = Single Anastomosis Duodeno-Ileal Bypass with Sleeve Gastrectomy; Band = Adjustable Gastric Band; BPD/DS = Biliopancreatic Diversion with Duodenal Switch; DML = Double Machine Learning; RF = Random Forest.

| Causal Method | **RYGB**<br>(vs. Sleeve) | **Band**<br>(vs. Sleeve) | **BPD/DS**<br>(vs. Sleeve) | **SADI-S**<br>(vs. Sleeve) |
| --- | --- | --- | --- | --- |
| Causal Forest | -0.0281<br>(-0.0483, -0.0079) | -0.7975<br>(-1.0182, -0.5768) | 0.2537<br>(0.1734, 0.3340) | -0.5198<br>(-0.6814, -0.3582) |
| Dragonnet | -0.0067<br>(-0.0076, -0.0058) | -0.9555<br>(-0.9572, -0.9539) | 0.2639<br>(0.2625, 0.2653) | -0.4294<br>(-0.4311, -0.4278) |
| DML (RF) | -0.0237<br>(-0.0361, -0.0113) | -0.7329<br>(-0.8070, -0.6588) | 0.2945<br>(0.2313, 0.3576) | -0.4107<br>(-0.4623, -0.3591) |
| DML (LightGBM) | -0.0268<br>(-0.0392, -0.0144) | -0.8150<br>(-0.8913, -0.7387) | 0.2867<br>(0.2208, 0.3526) | -0.4669<br>(-0.5144, -0.4194) |
| DML (XGBoost) | -0.0268<br>(-0.0392, -0.0144) | -0.8180<br>(-0.8937, -0.7423) | 0.2817<br>(0.2160, 0.3475) | -0.4445<br>(-0.4926, -0.3964) |

RYGB shows small negative ATEs ranging from −0.0067 to −0.0281, with confidence intervals consistently excluding zero, indicating a slightly lower 30-day reduction in BMI compared to Sleeve. Band demonstrates substantially negative ATEs in all methods, ranging from −0.7329 to −0.9555, indicating a significantly lower reduction in BMI within 30 days. BPD/DS consistently produces positive ATEs ranging from 0.2537 to 0.2945, indicating a significantly greater reduction in BMI than Sleeve within 30 days. SADI-S yields moderately negative ATEs ranging from *−*0.4107 to *−*0.5198, indicating a reduced short-term reduction in BMI compared to Sleeve alone.

### 3.3 Causal Risk Ratio on Reoperation, Readmission, and Intervention

Estimates of the causal risk ratio (RR) comparing each bariatric procedure to Sleeve are presented in Tables 3, 4, and 5 for reoperation, readmission, and intervention complications, respectively. These causal risk ratios were derived from average treatment effect estimations by substituting group differences with group ratios. Once again, across all four contrasts (RYGB vs. Sleeve, Band vs. Sleeve, BPD/DS vs. Sleeve, and SADI-S vs. Sleeve) and all three complication cases, the five ATE estimation and inference approaches produced directionally consistent results, indicating that the estimated causal risk ratios are robust to the choice among the considered machine-learning-based causal inference methods.

**Table 3:**
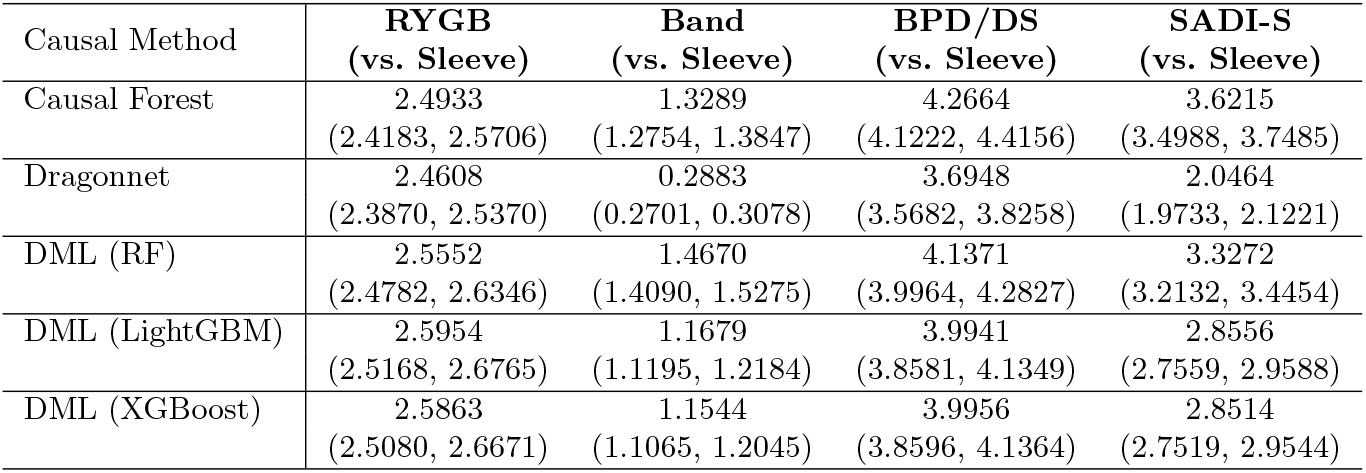
Causal risk ratios for 30-day reoperation comparing bariatric procedures to sleeve gastrectomy. Values represent risk ratios with 95% confidence intervals in parentheses. Risk ratios greater than 1.0 indicate higher reoperation rates compared to sleeve gastrectomy; values less than 1.0 indicate lower rates.

| Causal Method | **RYGB**<br>(vs. Sleeve) | **Band**<br>(vs. Sleeve) | **BPD/DS**<br>(vs. Sleeve) | **SADI-S**<br>(vs. Sleeve) |
| --- | --- | --- | --- | --- |
| Causal Forest | 2.4933<br>(2.4183, 2.5706) | 1.3289<br>(1.2754, 1.3847) | 4.2664<br>(4.1222, 4.4156) | 3.6215<br>(3.4988, 3.7485) |
| Dragonnet | 2.4608<br>(2.3870, 2.5370) | 0.2883<br>(0.2701, 0.3078) | 3.6948<br>(3.5682, 3.8258) | 2.0464<br>(1.9733, 2.1221) |
| DML (RF) | 2.5552<br>(2.4782, 2.6346) | 1.4670<br>(1.4090, 1.5275) | 4.1371<br>(3.9964, 4.2827) | 3.3272<br>(3.2132, 3.4454) |
| DML (LightGBM) | 2.5954<br>(2.5168, 2.6765) | 1.1679<br>(1.1195, 1.2184) | 3.9941<br>(3.8581, 4.1349) | 2.8556<br>(2.7559, 2.9588) |
| DML (XGBoost) | 2.5863<br>(2.5080, 2.6671) | 1.1544<br>(1.1065, 1.2045) | 3.9956<br>(3.8596, 4.1364) | 2.8514<br>(2.7519, 2.9544) |

**Table 4:** Causal risk ratios for 30-day readmission comparing bariatric procedures to sleeve gastrectomy. Values represent risk ratios with 95% confidence intervals in parentheses. Risk ratios greater than 1.0 indicate higher readmission rates compared to sleeve gastrectomy; values less than 1.0 indicate lower rates.

| Causal Method | **RYGB**<br>**(vs. Sleeve)** | **Band**<br>**(vs. Sleeve)** | **BPD/DS**<br>**(vs. Sleeve)** | **SADI-S**<br>**(vs. Sleeve)** |
| --- | --- | --- | --- | --- |
| Causal Forest | 1.8886<br>(1.8577, 1.9201) | 0.9602<br>(0.9385, 0.9825) | 2.5371<br>(2.4899, 2.5852) | 2.0026<br>(1.9644, 2.0417) |
| Dragonnet | 2.2236<br>(2.1867, 2.2610) | 0.6850<br>(0.6680, 0.7024) | 2.2020<br>(2.1602, 2.2447) | 3.0944<br>(3.0380, 3.1518) |
| DML (RF) | 1.9122<br>(1.8809, 1.9441) | 1.0309<br>(1.0080, 1.0544) | 2.5658<br>(2.5181, 2.6145) | 1.8752<br>(1.8389, 1.9123) |
| DML (LightGBM) | 1.9297<br>(1.8979, 1.9619) | 0.7712<br>(0.7527, 0.7902) | 2.3472<br>(2.3031, 2.3923) | 1.6631<br>(1.6302, 1.6967) |
| DML (XGBoost) | 1.9240<br>(1.8923, 1.9561) | 0.7772<br>(0.7585, 0.7962) | 2.3367<br>(2.2927, 2.3816) | 1.6668<br>(1.6339, 1.7004) |

**Table 5:** Causal risk ratios for 30-day intervention comparing bariatric procedures to sleeve gastrectomy. Values represent risk ratios with 95% confidence intervals in parentheses. Risk ratios greater than 1.0 indicate higher intervention rates compared to sleeve gastrectomy; values less than 1.0 indicate lower rates.

| Causal Method | **RYGB**<br>**(vs. Sleeve)** | **Band**<br>**(vs. Sleeve)** | **BPD/DS**<br>**(vs. Sleeve)** | **SADI-S**<br>**(vs. Sleeve)** |
| --- | --- | --- | --- | --- |
| Causal Forest | 2.6902<br>(2.6101, 2.7727) | 1.0781<br>(1.0326, 1.1256) | 3.4774<br>(3.3574, 3.6017) | 3.8206<br>(3.6920, 3.9536) |
| Dragonnet | 1.5190<br>(1.4752, 1.5641) | 0.2840<br>(0.2659, 0.3033) | 6.6842<br>(6.4593, 6.9169) | 2.5138<br>(2.4266, 2.6042) |
| DML (RF) | 2.7394<br>(2.6579, 2.8235) | 1.3322<br>(1.2785, 1.3882) | 3.1599<br>(3.0495, 3.2743) | 3.8897<br>(3.7586, 4.0254) |
| DML (LightGBM) | 2.8226<br>(2.7382, 2.9096) | 1.0491<br>(1.0046, 1.0956) | 3.0356<br>(2.9292, 3.1459) | 3.6336<br>(3.5106, 3.7610) |
| DML (XGBoost) | 2.8296<br>(2.7449, 2.9168) | 1.0488<br>(1.0043, 1.0953) | 3.0613<br>(2.9540, 3.1724) | 3.6713<br>(3.5473, 3.7996) |

RYGB demonstrates elevated risk ratios in all complication categories compared to Sleeve. Reoperation RRs range from 2.46 to 2.60, readmission RRs from 1.89 to 2.22, and intervention RRs from 1.52 to 2.83. The Band has the most favorable risk profile across all complication categories. Reoperation RRs range from 0.29 to 1.47, readmission RRs from 0.69 to 1.03, and intervention RRs from 0.28 to 1.33, with many estimates below 1.0 suggesting a potentially protective effect compared to Sleeve. BPD/DS demonstrates the highest overall risk ratios. Reoperation RRs range from 3.69 to 4.27, readmission RRs from 2.20 to 2.57, and intervention RRs show the most significant variability (3.04-6.68). SADI-S shows moderate to substantially elevated risk ratios, with considerable variation across estimation methods. Reoperation RRs range from 2.05 to 3.62, readmission RRs show the greatest variability (1.66-3.09), and intervention RRs are consistently high (2.51-3.89).

## 4 Discussion

Our analysis reveals substantial differences in short-term BMI reduction and 30-day complication rates across procedures, with important implications for surgical decision-making. Consistency of effect directions across all estimation methods (Causal Forest, Dragonnet, and Double Machine Learning with multiple base learners) strengthens confidence in these findings. Our causal estimates, derived using machine learning methods that flexibly adjust for nonlinear confounding, provide clinically actionable effect sizes quantifying what would happen if similar patients received different procedures–precisely the counterfactual question clinicians face. Concordance with mechanistic understanding from prior RCTs and physiological principles provides external validation.

### 4.1 Interpretation of BMI Reduction Findings

#### Roux-en-Y Gastric Bypass (RYGB) vs. Sleeve

The small negative ATEs for RYGB (−0.0067 to −0.0281) align with the physiological design of this procedure, which, although it reroutes the small intestine to promote long-term weight loss through malabsorption, demonstrates a comparable short-term impact to the more straightforward sleeve procedure [Manning et al., 2015]. While RYGB shows superior long-term weight loss outcomes [Arterburn et al., 2018], studies consistently demonstrate that sleeve gastrectomy achieves equivalent or slightly better short-term weight loss, with the comparative advantage of RYGB emerging primarily after the first postoperative year when the malabsorption mechanisms become more pronounced.

#### Adjustable Gastric Band (AGB, or Band) vs. Sleeve

The substantially negative ATEs for Band (−0.7329 to −0.9555) align with the minimally invasive approach of this procedure, which places an adjustable band around the proximal stomach to restrict food intake. While this restrictive mechanism produces lower complication rates and maintains excellent adjustability [Snyder et al., 2010], it inherently results in slower and more gradual weight loss trajectories compared to resectional procedures [Hady et al., 2012]. The pronounced negative effect reflects the fundamental mechanistic difference: Band relies solely on mechanical restriction and behavioral modification, requiring patients to adjust eating patterns, while Sleeve Gastrectomy involves both restriction and hormonal changes that promote greater immediate weight loss through ghrelin reduction and altered gastric emptying.

#### Biliopancreatic Diversion with Duodenal Switch (BPD/DS) vs. Sleeve

The substantial positive effects for BPD/DS (0.2537 to 0.2945) reflect its unique combination of gastric restriction through sleeve gastrectomy with extensive malabsorptive bypass, creating immediate anatomical changes and rapid onset of malabsorption [Anderson et al., 2013]. The aggressive weight loss profile is well documented, with studies demonstrating superior short-term results compared to purely restrictive procedures [Sucandy et al., 2014], confirming its efficacy despite greater surgical complexity and perioperative risk [Hutter et al., 2013]. The positive ATE demonstrates that the immediate metabolic effects of BPD/DS, including dramatic reduction in fundectomy-mediated ghrelin production and rapid nutrient diversion, translate into measurably superior early weight loss compared to Sleeve Gastrectomy.

#### Single Anastomosis Duodeno-Ileal Bypass with Sleeve Gastrectomy (SADI-S) vs. Sleeve

The moderately negative ATEs for SADI-S (−0.4107 to −0.5198) align with the design of this procedure, where the restrictive component drives most of the weight loss during the initial period. Recent evidence suggests that the restrictive component drives most weight loss and metabolic adaptations during the first 12 months, with weight trajectories and metabolic profiles not differing significantly from sleeve gastrectomy alone during this early period [Pereira et al., 2024]. The moderately negative ATE provides quantitative evidence of the early performance profile of SADI-S, confirming that its primary benefits are intended for long-term metabolic improvement rather than immediate weight loss enhancement, supporting the clinical rationale for this surgical approach.

### 4.2 Interpretation of Complication Risk Findings

#### Roux-en-Y Gastric Bypass (RYGB) vs. Sleeve

The elevated risk ratios for RYGB across all complication categories are attributable to the inherent gastrojejunal anastomosis, making it more surgically complex than single anastomosis procedures [Fringeli et al., 2015]. The dual anastomosis design introduces multiple potential failure points, contributing to a moderate but consistent elevation of risk in all evaluated complications.

#### Adjustable Gastric Band (AGB, or Band) vs. Sleeve

The favorable risk profile for Band is attributed to the preservation of normal digestive anatomy during the procedure, as it does not involve gastric resection or intestinal rerouting, thus minimizing immediate surgical trauma [Kodner and Hartman, 2014]. The absence of anastomoses further eliminates the acute leak risks characteristic of more complex procedures.

#### Biliopancreatic Diversion with Duodenal Switch (BPD/DS) vs. Sleeve

The substantially elevated risk for BPD/DS is consistent with the medical literature describing this procedure as requiring sleeve gastrectomy and duodenal division with extensive intestinal rearrangement, creating numerous potential failure points compared to single-procedure operations [Pereira et al., 2021]. The inherent technical complexity and extensive anatomical reconstruction account for its classification as the highest-risk procedure in most complication categories.

#### Single Anastomosis Duodeno-Ileal Bypass with Sleeve Gastrectomy (SADI-S) vs. Sleeve

The elevated risk profile for SADI-S is consistent with evidence suggesting that this procedure carries more complications than RYGB and Sleeve Gastrectomy, although comparable to BPD/DS. The current early experience with SADI-S may correlate with its observed complication profile [Clapp et al., 2023], with the learning curve associated with this newer procedure likely contributing to the increased risk of complications.

## 5 Conclusion

This large-scale causal analysis provides robust evidence that bariatric surgery procedures differ substantially in their balance between weight loss efficacy and safety, with BPD/DS achieving superior early BMI reduction at the cost of higher complication rates. At the same time, sleeve gastrectomy offers a favorable risk-benefit profile, supporting its current status as the most commonly performed procedure. The consistency of findings across multiple state-of-the-art causal inference methods strengthens confidence in these estimates. It demonstrates the value of applying machine-learning-based approaches to observational data in surgical comparative effectiveness research. By providing the first rigorous causal estimates of treatment effects across bariatric procedures, our findings move beyond prior associational evidence to enable truly evidence-based surgical decision-making. These quantitative causal estimates allow clinicians and patients to weigh the trade-offs between efficacy and safety based on patient-specific metabolic disease severity, comorbidity burden, and risk tolerance–transforming surgical selection from pattern-based associations to causal understanding of treatment effects.

## Data Availability

The dataset is publicly available

https://www.facs.org/quality-programs/accreditation-and-verification/metabolic-and-bariatric-surgery-accreditation-and-quality-improvement-program/participant-use-data-file-puf/

## Acknowledgements

We are profoundly grateful to Professor Carlos Fernandez-Granda (Courant Institute of Mathematical Sciences and Center for Data Science, New York University, New York, NY) for his instrumental role in shaping this research. As the undergraduate advisor to the first author, Professor Fernandez-Granda provided invaluable guidance, mentorship, and critical insights throughout the conceptualization and early development of this project. His generous support and thoughtful direction were fundamental to the success of this work.

## Appendix

### A Complete List of Preoperative Covariates

The following preoperative variables from the MBSAQIP registry were included in all causal inference analyses, organized by category:

#### Demographic and Baseline Factors

- Age
- Sex
- Race
- Body Mass Index (BMI)
- Preoperative Functional Health Status
- Preoperative Is the Patient’s Ambulation Limited Most or all of the Time

#### Cardiovascular and Metabolic Comorbidities

- Preoperative Hypertension Requiring Medication
- Preoperative Number of Anti-Hypertensive Medications
- Preoperative Hyperlipidemia Requiring Medication
- Preoperative Diabetes Mellitus Requiring Therapy with Non-Insulin Agents or Insulin
- History of Myocardial Infarction
- Previous PCI/PTCA
- Previous Cardiac Surgery
- Preoperative Vein Thrombosis Requiring Therapy
- Preoperative Venous Stasis
- Preoperative Currently Requiring or On Dialysis
- Preoperative Renal Insufficiency

#### Respiratory Conditions

- History of Severe COPD
- Preoperative Oxygen Dependent
- History of Pulmonary Embolism
- Preoperative Obstructive Sleep Apnea Requiring CPAP/BiPAP (or similar technology)

#### Surgical History and Clinical Indicators

- Previous Obesity Surgery/Foregut Surgery
- Revision/Conversion Principal Operative Procedure
- Mini-Loop Gastric Bypass (MGB) Principal Operative Procedure
- Gastroesophageal Reflux Disease (GERD) Requiring Medication (within 30 days prior to surgery)
- Current Smoker within One Year
- Preoperative Steroid/Immunosuppressant Use for a Chronic Condition
- Preoperative Therapeutic Anticoagulation
- Preoperative Does the patient have an IVC filter
- ASA Classification Principal Operative Procedure
- Was a Swallow Study Performed the Day of or the Day After the Procedure

These covariates span demographic characteristics, cardiovascular and metabolic comorbidities, respiratory conditions, and surgical history with clinical indicators.

### B Technical Details of Causal Inference Methods

#### Potential Outcome Framework

We employ the potential outcomes framework to estimate causal effects. All preoperative variables that can affect postoperative outcomes, except types of surgery, can be defined as *covariate, X*. We designated sleeve gastrectomy as the reference procedure and performed pairwise comparisons with other surgical types. For each comparison, *T* represents the *treatment* (*T* = 0 for sleeve gastrectomy, *T* = 1 for the comparator procedure). The *propensity score* is defined as P(*T* = 1|*X* = *x*), representing the probability of receiving the comparator treatment given covariates *X*. The *potential outcomes* are denoted as *Y ∈* {*Y*_1_, *Y*_0_}, with the outcome models *µ*_1_(*x*) and *µ*_0_(*x*) specified as *Y*_*t*_(*x*) = *µ*_*t*_(*x*) + *E*_*t*_ for *t* = 1, 0. For continuous results (BMI reduction), we estimate *average treatment effect (ATE)*.

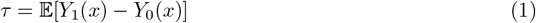

Positive ATE values indicated superior effectiveness of the comparator procedure compared to sleeve gastrectomy. For binary outcomes (complications), we estimate the *causal risk ratio (RR)*.

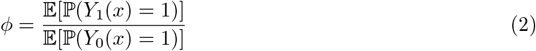

Risk ratios greater than 1.0 indicated higher complication rates for the comparator procedure versus sleeve gastrectomy.

#### B.2 Assumptions

Our causal inference approach relies on three fundamental assumptions:

- **Stable Unit Treatment Value Assumption (SUTVA):** The potential outcomes for any unit do not vary with the treatments assigned to other units, and, for each unit, there are no different forms or versions of each treatment level, which lead to various potential outcomes. In our study, this means the outcomes for each patient are not affected by the treatment assignment of other patients.
- **Unconfoundedness Assumption:** The potential outcomes are independent of the assignment of the treatment, given the covariates, i.e. (*Y*_1_, *Y*_0_) ⊥ *T* | *X*. This assumption posits that all variables affecting both treatment selection and outcomes are measured and included as covariates. While randomized controlled trials would eliminate this concern through randomization, ethical and practical constraints make RCTs infeasible for comparing established surgical procedures.
- **Overlap Assumption:** There is a positive probability of receiving both treatment and control for all relevant covariate values, i.e. *δ* ≤P(*T* = 1 |*X* = *x*) ≤ 1 − *δ* for 0 < *δ* < 1. This assumption ensures that every patient type represented in the data has a nonzero probability of receiving either procedure, allowing for meaningful counterfactual comparisons.

#### B.3 Causal Inference Methods

Under the above assumptions, we implemented three complementary causal inference approaches:

- **Causal Forest:** This method uses tree-based ensemble learning to estimate heterogeneous treatment effects across patient subgroups. The algorithm constructs a forest of causal trees, where each tree is built using a modified splitting criterion that maximizes treatment effect heterogeneity rather than outcome prediction accuracy. The final treatment effect estimate combines predictions across trees using inverse propensity weighting to reduce selection bias. Honest inference is maintained by using separate subsamples for tree construction and effect estimation.
- **Double Machine Learning (DML):** This framework employs a two-stage cross-fitting procedure to separately estimate the propensity score and outcome models using flexible machine learning algorithms. We implemented DML with three different base learners: Random Forest, LightGBM, and XGBoost. The method reduces bias from model misspecification by orthogonalizing (removing confounding between) treatment assignment and outcomes through residualization.
- **Dragonnet:** This neural network architecture jointly learns propensity score and outcome models through shared representations while applying targeted regularization to reduce selection bias. The network structure includes shared layers that learn a common representation of covariates, followed by separate heads for propensity estimation and outcome prediction. A specialized regularization term encourages the learned representations to balance treatment groups, reducing sensitivity to propensity score misspecification.

For binary complication outcomes, we adapted these methods to estimate risk ratios by replacing additive outcome differences with multiplicative outcome ratios in the final aggregation step.

